# The impact of frailty on the outcomes of COVID-19 patients with persistent critical illness: A population-based cohort study

**DOI:** 10.1101/2023.07.17.23292714

**Authors:** William Bonavia, Ravindranath Tiruvoipati, Mallikarjuna Ponnapa Reddy, David Pilcher, Ashwin Subramaniam

## Abstract

**Objectives:** Persistent critical illness (PerCI, ≥10 days in Intensive Care Unit [ICU]) is defined as the time from ICU admission when patients’ antecedent characteristics define their mortality rather than the admission aetiology. Patients with frailty and without COVID-19 have a higher risk of developing and dying from PerCI. We aimed to investigate the impact of frailty on critically ill patients with COVID-19 experiencing PerCI.

**Methods:** We conducted a retrospective multicentre cohort study including 103 Australian and New Zealand ICUs over two years, investigating the impact of frailty, measured with Clinical Frailty Scale (CFS), in patients with COVID-19, between patients with and without PerCI.

**Results:** The prevalence of PerCI was similar between patients with and without frailty (25.4% vs. 27.9%; p=0.44). Hospital mortality was higher in patients with PerCI than without (28.8% vs. 9.3%; p<0.001), with mortality rising with increasing CFS (p<0.001). Frailty independently predicted hospital mortality, but when adjusted for ANZROD and sex, its impact was no different in patients with and without PerCI (odds ratio [OR]=1.30 [95%-CI: 1.14-1.49] vs. OR=1.46 [95%-CI: 1.29-1.64]).

**Conclusions:** The presence of frailty independently predicted hospital mortality in patients with PerCI, but frailty did not have a different impact on patients with and without PerCI.

## Introduction

With an improved understanding of critical illness and the advent of new technologies, an increasing proportion of patients survive initial insult, only to require various supports necessitating intensive care for a prolonged period. In recent years, efforts have been made to better characterise critically ill patients requiring prolonged intensive care unit (ICU) admissions, to better understand the underlying causative factors, as well as outcomes, of this patient group. [1–4]

Persistent Critical Illness (PerCI) is a novel domain in ICU gaining clinical significance rapidly. PerCI is the point in a patient’s ICU admission in which the outcome is no longer driven by the aetiology of the admission but by the new array of complications they have suffered from a prolonged ICU stay. The driving factors, rather than the primary pathology, become the antecedent characteristics of the patient.[5,6] These antecedent characteristics are generally identified to be patient demographics, underlying comorbidities, prior living circumstances, and chronic factors from acute illness severity scores. Numerous large observational studies have been performed in multiple countries and have identified this point tends to occur between day 5 and day 22 (day 10 in Australian, New Zealand and North American populations, day 11 in UK population). [7–11] Approximately 5% to 35% of patients admitted to ICU develop PerCI. [7–11] Such patients generally have relatively poor clinical outcomes (mortality, longer hospitalisation) and functional outcomes (functional dependence, disability, and quality of life).[7,8,12,13]

Within the field of critical care, there is growing awareness of the frailty syndrome, a complex state of reduced physiologic reserve that is associated with but not necessarily present with senescence. [14] Previous studies have investigated the impact of frailty on critically ill patients, identifying both increased risk of mortality, and increased likelihood of discharge to a residential aged care facility.[15–19] A recent study identified that there was an increased likelihood of both developing and dying from PerCI in patients with frailty. [20] Patients with higher degrees of frailty have near double the risk of developing PerCI when compared with their less frail counterparts.[20] Furthermore, there is a progressive increase in mortality in patients with frailty throughout their ICU admission.[20]

Critically ill patients with COVID-19 frequently suffer from multiorgan failure and have prolonged ICU stays (mean duration of 10.8 days,[21] twice that of severe community-acquired pneumonia).[22] The impact of frailty on mortality in this group is unclear, as previous observational studies have demonstrated a mixed picture.[23,24]

Currently, there are no published studies from Australia or New Zealand that explore PerCI in patients with COVID-19. Given the continued prevalence of the COVID-19 pandemic with an ongoing need for intensive care resources, a better understanding of the outcomes in patients with frailty with COVID-19 that develop PerCI is needed. In this study, we aim to examine the impact of frailty on critically ill patients with COVID-19 experiencing PerCI.

## Methods

### Ethics approval

The study was approved by the Alfred Hospital Ethics Committee (Project No: 176/21). Access to ANZICS Adult Patient Database was granted by the ANZICS Centre for Outcome and Resource Evaluation Management Committee following standing protocols.

### Study Design and Setting

This was a retrospective multicentre cohort study, analysing the Australian and New Zealand Intensive Care Society (ANZICS) Adult Patient Database (APD) between 1^st^ January 2020 to 31^st^ December 2021.

### Patient Identification

All consecutive critically ill adult patients (age ≥16 years) with COVID-19 with documented clinical frailty scale (CFS) score and admitted to Australian and New Zealand ICUs with an Acute Physiology and Chronic Health Evaluation (APACHE) III-j admission diagnostic codes for viral pneumonia or acute respiratory distress syndrome (ARDS) were included. Patients with COVID-19 were identified using the ANZICS modification of the APACHE IV diagnostic system which codes the primary cause of ICU admission (Supplementary Table 1). Readmission episodes during the same hospitalisation and admissions for palliative care or potential organ donation were also excluded.

### ANZICS-APD

The ANZICS-APD contains routine quality assurance and benchmarking data collected by ANZICS CORE. This database includes de-identified patient data from more than 97% of Australian and more than 65% of New Zealand ICUs, including admission diagnosis, chronic health status, and physiological and biochemical variables within the first 24 hours of admission. The definitions of each condition are described in the ANZICS-APD data dictionary.[25]

### Definitions

For this study, we defined PerCI as patients with ICU length of stay of 10 days and longer.

### Frailty assessment

Frailty was identified using the Canadian Study of Health and Aging Clinical Frailty Scale (CFS). This nine-point tool quantifies frailty based on the deficit accumulation approach.[26] This scale ranges from CFS=1 (very fit), 2 (well), 3 (managing well), 4 (vulnerable), 5 (mildly frail), 6 (moderately frail), 7 (severely frail), 8 (very severely frail) to 9 (terminally ill). The CFS has been validated amongst critically ill patients [16,27] with good inter-rater reliability, [27,28] and has been correlated with the other frailty scales. [29,30] In the ANZICS-APD, the CFS is modified to eight categories without a CFS of 9 (terminally ill). [31] The CFS represented the patient’s status in the two months preceding ICU admission. [31] For this study, we further grouped CFS scores according to four groups, CFS-1-2, CFS-3-4, CFS-5-6, and CFS-7-8 as reported in a recent study. [20]

### Exposure and confounding variables

The exposure variable was frailty status based on CFS categories in patients with and without PerCI. The confounding variables were illness severity (measured with ANZROD), and sex.

### Study outcomes

We aimed to investigate whether the impact of frailty in COVID-19 patients differed in those with and without PerCI. The primary outcome was hospital mortality. Secondary outcomes included ICU mortality, ICU and hospital length of stays, resource burden (ICU Bed days and Hospital Bed days used), and discharge destination.

### Statistical Analysis

Normality was assessed in continuous data by employing the Shapiro-Wilk test. The group comparisons between patients with and without PerCI were made using chi-square tests for proportions, student t-tests for normally distributed data and Mann-Whitney U or Kruskal-Wallis tests for non-parametric data depending on the numbers of categories examined. Categorical data are reported as frequencies (%). Normally distributed data were reported using the mean (standard deviation [SD]) and non-parametric data were reported as medians (interquartile range [IQR]). Illness severity was determined using ANZROD, a highly discriminatory, locally derived, and well-calibrated mortality prediction model used for benchmarking ICU performance in ANZ which combines age, chronic illnesses, acute physiological disturbance, and diagnosis.[31,32] The association of CFS with hospital mortality in the PerCI and non-PerCI groups was investigated using multivariable logistic regression, for frail and non-frail patients, and the results reported as odds ratio (OR) and 95% confidence interval (CI). Kaplan-Meier survival curves were performed based on frailty status, both dichotomized and based on categories. Model discrimination was assessed using the area under the receiver operating characteristic (AUROC) plots with a comparison between models assessed using chi-square tests.[33] Analyses were performed using SPSS software (version 27), and a two-sided p-value of <0.05 was used to indicate statistical significance.

## Results

### Patient Characteristics

A total of 3722 patients with COVID-19 were admitted to 103 Australian and New Zealand ICUs during the study period with admission diagnoses of either viral pneumonia or ARDS reported to the ANZICS-APD. Of these, 3064 with a documented CFS were included in the final analysis (Supplementary Figure 1). The comparison of those with and without CFS values is summarised in Supplementary Table 2. Although there were some group differences, there were no differences in the illness severity scores between the two groups.

Baseline characteristics are presented in Table 1. Patients with PerCI were older (median [IQR] 59.6 [48.9-69.1] vs. 55.8 [42.0-68.1]; p<0.001) and more frequently male (66.0% vs. 59.5%; p=0.001), when compared with those without PerCI. Patients with PerCI were more likely to be transferred from another ICU (24.6% vs. 7.5%; p<0.001) than those without PerCI. Patients with PerCI less frequently had chronic comorbidities such as respiratory and cardiovascular conditions, or obesity, but more commonly had diabetes mellitus and delirium, than patients without PerCI. There was no difference in median [IQR] CFS or illness severity scores between the two groups. Patients that developed PerCI less frequently had treatment limitations at ICU admission (4.4% vs. 9.5%; p<0.001) than those without PerCI. More patients with PerCI needed ICU organ supports for mechanical and non-invasive ventilation, tracheostomies, and vasoactive, ECMO and renal replacement therapies than patients without PerCI. ICU discharge delay was longer in patients with PerCI, and the median (IQR) was lower than 6 hours in both groups (4.0 [2.0-7.3] vs. 4.7 [2.2-8.8]; p<0.001). Further categorisation by CFS categories is provided in Supplementary Tables 3a and 3b.

**Table 1:** Baseline characteristics of patients with and without persistent critical illness with COVID-19 pneumonitis.

| Variable | Patients without Persistent Critical illness (n=2217) | Patients with Persistent Critical illness (n=847) | p-value |
| --- | --- | --- | --- |
| Frailty status, n (%) |  |  |  |
| CFS - Frailty score (median [IQR]) | 3 (2, 4) | 3 (2, 3) | 0.29 |
| - CFS-1-2 | 835 (37.7%) | 288 (34.0%) | 0.06 |
| - CFS-3-4 | 1109 (50.0%) | 466 (55.0%) |  |
| - CFS-5-6 | 219 (9.9%) | 80 (9.4%) |  |
| - CFS-7-8 | 54 (2.2%) | 13 (1.5%) |  |
| Male sex, n (%) | 1320 (59.5%) | 559 (66.0%) | 0.001 |
| Indigenous status, n (%) | 61 (2.8%) | 46 (5.4%) | 0.50 |
| Age, (years), median (IQR) | 55.8 (42.0, 68.1) | 59.6 (48.9, 69.1) | <0.001 |
| Admission source, n (%) |  |  |  |
| - Home | 1891 (85.3%) | 582 (68.7%) | <0.001 |
| - Other acute hospital | 192 (8.7%) | 88 (4.6%) |  |
| - Nursing home or chronic care | 12 (0.5%) | 3 (0.4%) |  |
| - Other hospital ICU | 94 (4.2%) | 212 (25.0%) |  |
| - Rehabilitation | 3 (0.1%) | 0 (0) |  |
| - Missing | 25 (1.1%) | 11 (1.3%) |  |
| ICU admission source, n (%) |  |  |  |
| - Emergency Department (ED) | 937 (42.3%) | 254 (30.0%) | <0.001 |
| - Ward | 1103 (49.8%) | 380 (44.9%) |  |
| - Other hospital (ED and ICU) | 156 (7.5%) | 209 (24.6%) |  |
| - Operating Theatre / Recovery | 1 (0.0%) | 0 (0) |  |
| - Direct admit | 10 (0.5%) | 4 (0.5%) |  |
| Documented co-morbidities, n (%) |  |  |  |
| - Chronic respiratory condition | 163 (7.4%) | 37 (4.4%) | 0.003 |
| - Chronic cardiovascular condition | 149 (6.7%) | 30 (3.5%) | <0.001 |
| - Chronic renal failure | 61 (2.8%) | 13 (1.5%) | 0.05 |
| - Chronic liver disease | 14 (0.6%) | 7 (0.8%) | 0.56 |
| - Diabetes mellitus | 632 (29.6%) | 279 (34.4%) | 0.013 |
| - Immune suppressive therapy | 100 (4.5%) | 42 (5.0%) | 0.60 |
| - Lymphoma | 8 (0.4%) | 5 (0.6%) | 0.38 |
| - Leukaemia | 18 (0.8%) | 8 (0.9%) | 0.72 |
| - Metastatic cancer | 20 (0.9%) | 4 (0.9%) | 0.23 |
| - Obese (BMI ≥ 30 kg.m<sup>-2</sup>) | 984 (44.4%) | 300 (35.4%) | <0.001 |
| Organ failure scores |  |  |  |
| - APACHE III (mean [SD]) | 47.5 (19.8) | 56.6 (18.8) | 0.10 |
| - ANZROD (%) (mean [SD]) | 9.0 (12.5) | 11.0 (11.4) | 0.93 |
| Miscellaneous |  |  |  |
| Delirium in ICU | 89 (4.0%) | 170 (20.1%) | <0.001 |
| Pregnancy status | 62 (2.8%) | 10 (1.2%) | <0.001 |
| Pre-ICU (days) (median (IQR)) | 14.1 (4.8, 58.6) | 8.0 (0.61, 52.1) | <0.001 |
| ICU Discharge delay, (median (IQR)) | 4.0 (2.0, 7.3) | 4.7 (2.2, 8.8) | <0.001 |
| ICU admission post MET call | 823 (37.4%) | 282 (33.7%) | 0.06 |
| Treatment limitations at ICU admission | 210 (9.5%) | 37 (4.4%) | <0.001 |
| Cardiac arrest, n (%) | 5 (0.2%) | 1 (0.1%) | 0.08 |
| ICU Supports |  |  |  |
| Mechanical ventilation (MV), n (%) | 584 (26.7%) | 722 (85.5%) | <0.001 |
| Non-invasive ventilation (NIV), n (%) | 875 (40.1%) | 385 (46.2%) | 0.002 |
| Vasopressor and inotropes, n (%) | 534 (24.4%) | 654 (77.7%) | <0.001 |
| Renal replacement therapy, n (%) | 45 (2.1%) | 135 (16.2%) | <0.001 |
| ECMO, n (%) | 12 (0.6%) | 92 (11.0%) | <0.001 |
| Tracheostomy, n (%) | 8 (0.4%) | 178 (21.5%) | <0.001 |
| CFS – clinical frailty scale, SD – standard deviation, IQR. – interquartile range, BMI – body mass index, MET – medical emergency team, APACHE - Acute Physiology and Chronic Health Evaluation, ED – emergency department, ICU – intensive care unit, ANZROD – Australia New Zealand risk of death. |  |  |  |

### Primary Outcome

Overall hospital mortality was higher in patients with PerCI (28.8% [239/847] vs. 9.3% [202/2,217]; p<0.001) than those without PerCI. Higher hospital mortality was observed in PerCI patients compared to those without PerCI at equivalent frailty categorical levels (p<0.001; Figure 1). The Kaplan-Meier survival curve categorised based on PerCI and frailty status is illustrated in Figure 2. After adjusting for baseline illness severity (ANZROD) and sex, frailty independently predicted hospital mortality, but the impact of frailty, adjusted for ANZROD and sex, was no different in patients with and without PerCI (Figure 1, Supplementary Table 4). This finding was also confirmed in a sensitivity analysis conducted using the CFS as a continuous variable (OR=1.30 [95%-CI: 1.14-1.49] vs. OR=1.46 [95%-CI: 1.29-1.64]) (Supplementary Table 4).

**Figure 1:**
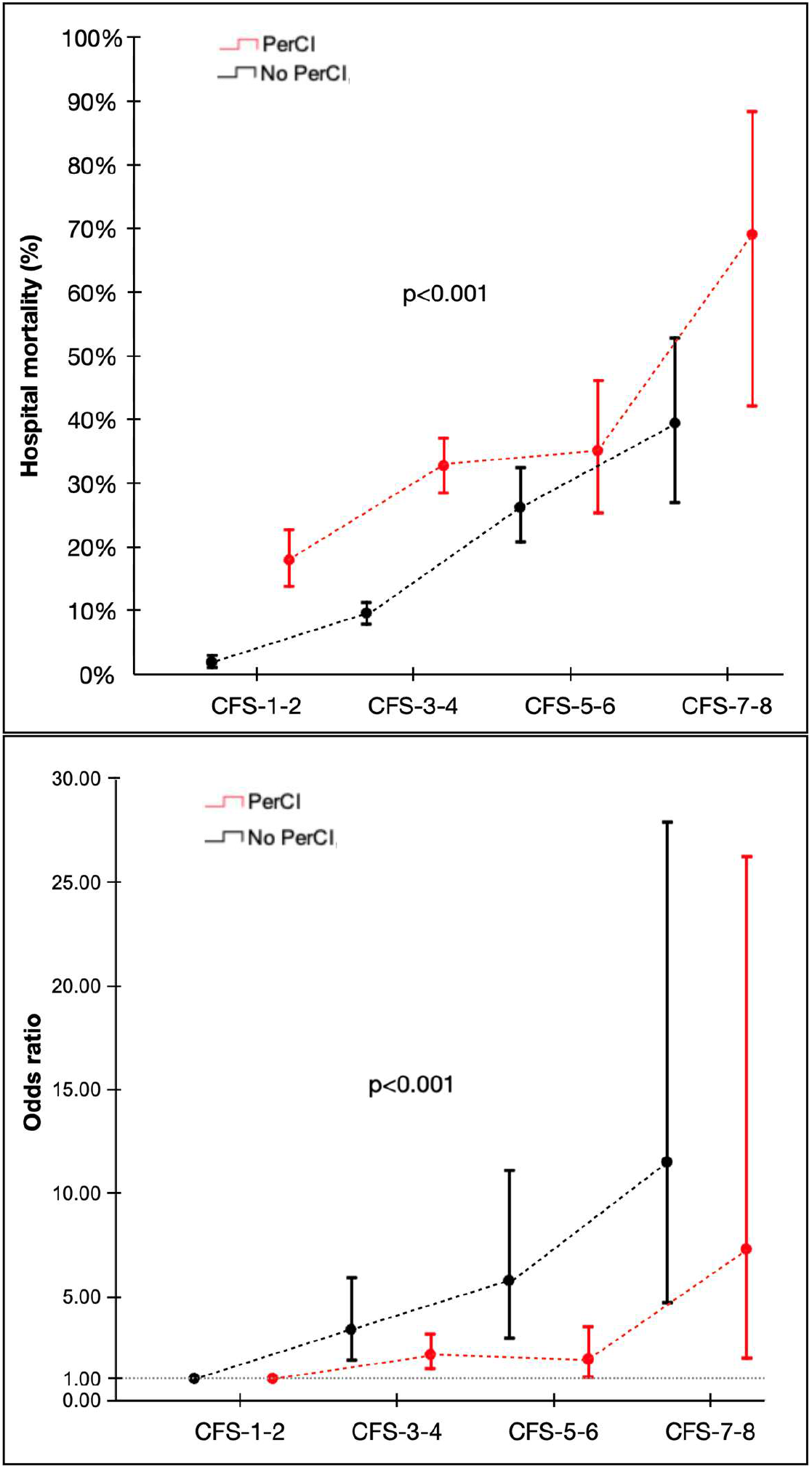
Hospital mortality according to Clinical Frailty Scale (CFS) score for all patients with and without PerCI. The top panel is unadjusted for hospital mortality, while the bottom panel is adjusted for ANZROD and sex. Please also refer to Supplementary Table 4.

**Figure 2:**
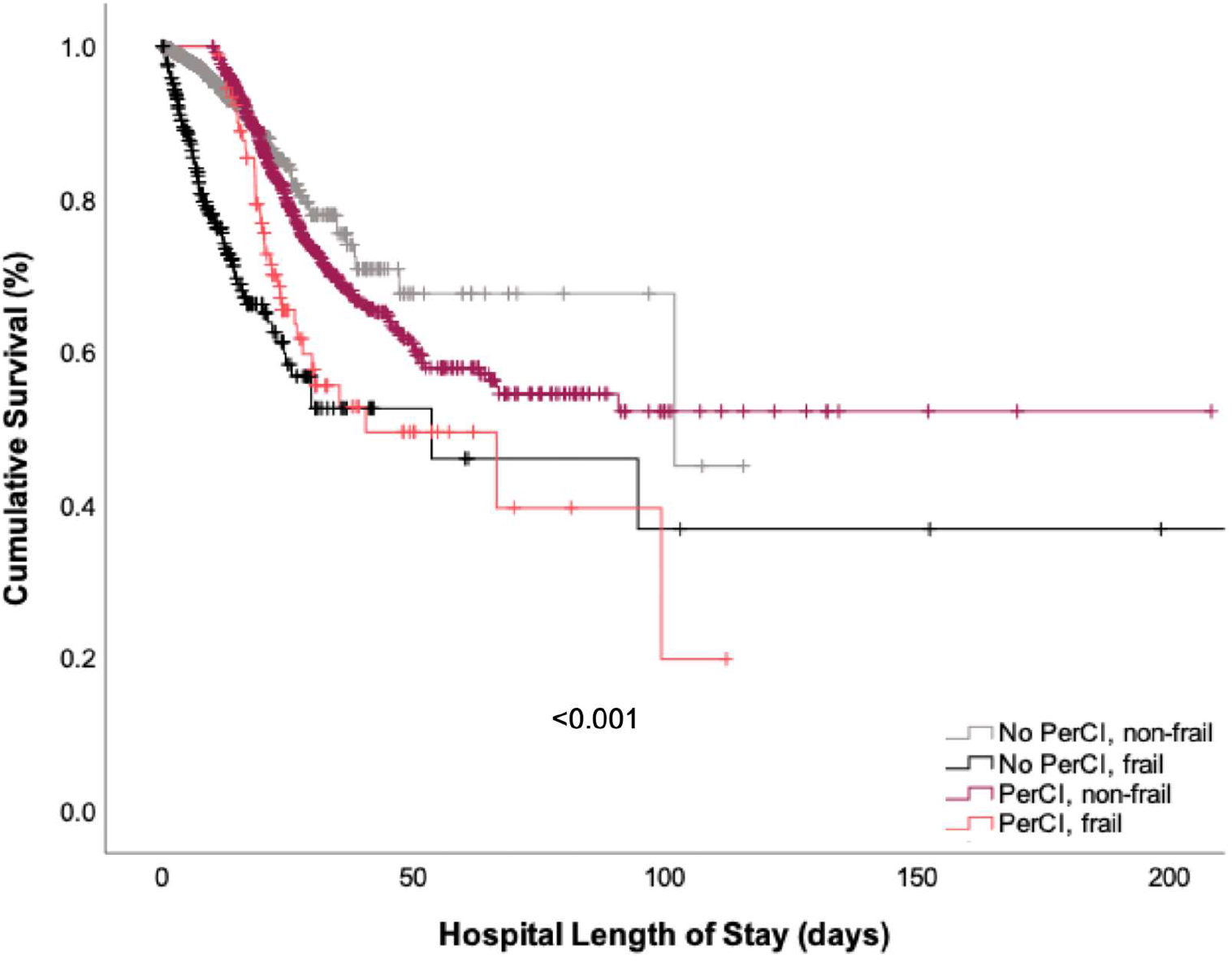
Kaplan Meier Survival Curve, stratified by PerCI and frailty status.

Frailty alone as a predictor of mortality showed only moderate discrimination in differentiating survivors from those who died. This effect was lower in patients with PerCI (AUROC 0.61 vs 0.76; p<0.001). Furthermore, the relationship between frailty and mortality in those with and without PerCI, before (AUROC 0.61 vs 0.76; p<0.001) and after adjusting for ANZROD and male sex (AUROC 0.70 vs 0.89; p<0.001) demonstrated that the mortality prediction was inferior in patients with PerCI (Supplementary Figure 2).

### Secondary Outcomes

The overall unadjusted ICU mortality rate was higher for patients with PerCI (25.6% vs. 5.9%; p<0.001), compared to patients without PerCI (Table 2). Patients with PerCI had a longer median length of stay in ICU than patients without PerCI (16.8 [IQR 12.7-25.7] vs. 5.0 [IQR 2.1-10.9] days; p<0.001; Table 2, Supplementary Figure 3). The median hospital length of stay was longer for patients with PerCI (25.1 [IQR 18.7-38.7] vs. 9.6 [IQR 6.0-14.9] days; p<0.001) than those without PerCI. Patients with frailty accounted for <10% of all ICU and hospital bed days, compared with patients without frailty (Figure 3). Overall, the patients with PerCI were less likely to be discharged home, when compared to patients without PerCI (p<0.001). The ICU readmissions and discharge to nursing homes were no different for both groups.

**Figure 3:**
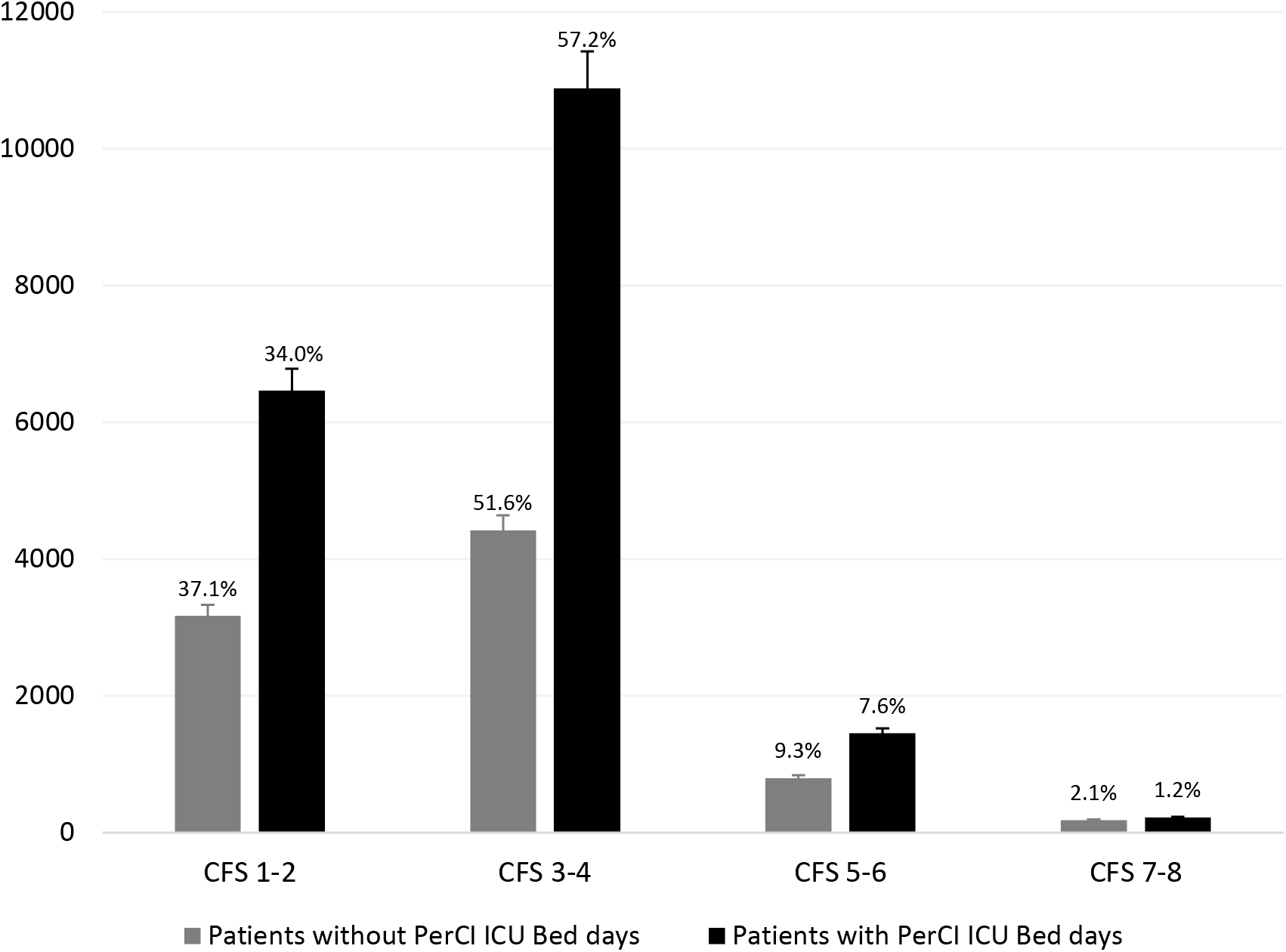
Total ICU bed days occupied stratified by CFS categories demonstrating that >90% of ICU bed-days for patients with PerCI were occupied by non-frail patients (CFS categories 1-2 and 3-4; p<0.001). *Error bars are standard errors of the mean.

**Table 2:** Unadjusted primary and secondary outcomes between patients with and without persistent critical illness, for the whole population and stratified by clinical frailty scale (CFS).

| Variable | Patients without Persistent Critical illness | Patients with Persistent Critical illness | p-value |
| --- | --- | --- | --- |
| Hospital mortality overall, n (%) | 202 (9.3%) | 239 (28.8%) | <0.001 |
| Hospital mortality by CFS categories, n (%) |  |  | <0.001 |
| - CFS-1-2 | 17/812 (2.1%) | 51/281 (18.1%) |  |
| - CFS-3-4 | 107/1094 (9.8%) | 151/458 (33.0%) |  |
| - CFS-5-6 | 57/215 (26.5%) | 28/79 (35.4%) |  |
| - CFS-7-8 | 21/53 (39.6%) | 9/13 (69.2%) |  |
| ICU mortality overall, n (%) | 131 (5.9%) | 217 (25.6%) | <0.001 |
| ICU mortality by CFS categories, n (%) |  |  | <0.001 |
| - CFS-1-2 | 10/835 (1.2%) | 47/288 (16.3%) |  |
| - CFS-3-4 | 71/1109 (6.4%) | 139/466 (29.8%) |  |
| - CFS-5-6 | 37/219 (16.9%) | 23/80 (28.7%) |  |
| - CFS-7-8 | 13/54 (24.1%) | 8/13 (61.5%) |  |
| ICU length of stay overall, median (IQR) | 5.0 (2.1, 10.9) | 16.8 (12.7, 25.7) | <0.001 |
| ICU length of stay by CFS categories, n (%) |  |  | <0.001 |
| - CFS-1-2 | 3.2 (1.7, 5.6) | 16.4 (12.5, 23.8) |  |
| - CFS-3-4 | 3.3 (1.8, 6.0) | 17.6 (13.1, 27.4) |  |
| - CFS-5-6 | 2.9 (1.3, 5.7) | 14.8 (12.1, 21.9) |  |
| - CFS-7-8 | 3.0 (1.5, 5.2) | 13.6 (11.6, 19.5) |  |
| Hospital length of stay overall, median (IQR) | 9.6 (6.0, 14.9) | 25.1 (18.7, 38.7) | <0.001 |
| Hospital length of stay by CFS categories, median (IQR) |  |  | <0.001 |
| - CFS-1-2 | 9.3 (6.0, 14.2) | 24.0 (18.2, 37.5) |  |
| - CFS-3-4 | 9.8 (6.0, 14.9) | 27.2 (19.3, 41.6) |  |
| - CFS-5-6 | 10.7 (5.4, 17.5) | 23.6 (18.7, 32.6) |  |
| - CFS-7-8 | 8.4 (4.4, 19.9) | 18.7 (15.2, 32.7) |  |
| ICU Readmission overall, n (%) | 101 (4.6%) | 26 (3.1%) | 0.07 |
| ICU Readmission by CFS categories, n (%) |  |  | 0.11 |
| - CFS-1-2 | 29/835 (3.5%) | 4/288 (1.4%) |  |
| - CFS-3-4 | 56/1109 (5.0%) | 18/466 (3.9%) |  |
| - CFS-5-6 | 12/219 (5.5%) | 3/80 (3.8%) |  |
| - CFS-7-8 | 7/54 (7.4%) | 1/13 (7.7%) |  |
| Home discharge overall, n (%) | 1541 (69.5%) | 313 (37.1%) | <0.001 |
| Home discharge by CFS categories, n (%) |  |  | <0.001 |
| - CFS-1-2 | 675/835 (80.8%) | 130/288 (45.1%) |  |
| - CFS-3-4 | 744/1109 (67.1%) | 149/466 (32.0%) |  |
| - CFS-5-6 | 105/219 (47.9%) | 33/80 (41.3%) |  |
| - CFS-7-8 | 17/54 (31.5%) | 1/13 (7.7%) |  |
| New Nursing home discharge overall, n (%) | 17 (0.8%) | 7 (0.8%) | 0.87 |
| New Nursing home discharge by CFS categories, n (%) |  |  | 0.32 |
| - CFS-1-2 | 0/835 (0) | 3/288 (1.0%) |  |
| - CFS-3-4 | 6/1109 (0.5%) | 1/466 (0.2%) |  |
| - CFS-5-6 | 7/219 (3.2%) | 3/80 (3.8%) |  |
| - CFS-7-8 | 4/54 (7.4%) | 0/13 (0) |  |

## Discussion

In this multicentre retrospective observational study that investigated patients with and without PerCI, admitted to ICU in Australia and New Zealand, we hypothesized that the presence of frailty would impact patient outcomes. Although patients with PerCI were younger, had fewer comorbidities and were less likely to have treatment limitations at ICU admission, there was no difference in the frailty status between the two groups. Hospital mortality was higher in patients with PerCI, but the impact of frailty was similar in patients with and without PerCI. Finally, frailty independently predicted hospital mortality in both patients with and without PerCI.

### Comparison to Published Literature

Although vaccination may have minimized the severity and mortality of the disease, many patients are still affected by the consequences of COVID-19 and develop PerCI. Although some studies have investigated the outcomes of patients either chronically or persistently critically ill with COVID-19, [34–37] there has been no exploration to date investigating the interplay between frailty and PerCI in COVID-19. A large study early in the pandemic found that almost half of all patients with COVID-19 admitted to critical care developed PerCI, with high resource use in critical care and beyond. [37] However, patients who developed PerCI were not associated with an increase in overall mortality when compared with patients that did not develop PerCI. [37] Mortality was analysed at 1 year, potentially accounting for this difference in outcomes. [37]

A recent study from Australia and New Zealand that explored the impact of frailty on patients without COVID-19 with PerCI, found that frailty resulted in an increased risk in hospital mortality. [20] Additionally the authors observed that frailty acted as a negative prognostic factor, with an increased risk of death over time throughout their hospital stay. [20] In contrast, our study found that hospital mortality increased with increasing frailty, but most patients with frailty with COVID-19 died early, frailty thereby losing predictive potency over time. Our study also found that patients with frailty accounted for a very small fraction of the ICU bed days. This is in keeping with previous studies that had observed similar findings.[23,24,38] Although resource constraints could be speculated for the shorter ICU stay in patients with frailty, a recent study from Australia and New Zealand found that patients were treated on merit and there was no difference in care between patients with and without COVID-19. [38] Additionally, the heterogeneity of the COVID-19 disease with varying rates of disease outcomes over the course of the pandemic could have played a role. [39]

Previous studies identified that developing PerCI substantially reduced the chances of discharge directly home from the hospital.[7] This was consistent with our findings, where increasing degrees of frailty resulted in an incrementally reduced chance of discharge home, in patients who developed PerCI, as well as those that did not. A recent study found that ‘long-COVID’ frequented in patients with severe COVID-19 and those with prolonged hospital stay. [40] Although PerCI describes a statistical outcome representative of prolonged disease state, we can contemplate that many of the consequences of PerCI can persist in the longer term (‘long-COVID’), [41] and potentially affect the cardiovascular, musculoskeletal, and neurologic systems.[41–43]

### Study Implications

It is known that patients admitted to ICU that develop PerCI have higher hospital mortality than those that do not develop PerCI, and this holds true in the novel disease process that is COVID-19. However, although frailty represents an independent predictor of hospital mortality, its impact is not distinctly different when contrasting patients with and without PerCI. Therefore, cautious, individualized, evidence-informed patient-centred care for patients with frailty and critical COVID-19 is essential. Conversations about goals of care (including advanced care planning, and therapeutic limitations) in the context of severe COVID-19 may be essential for those with multiple chronic conditions and greater baseline frailty levels (CFS ≥4). [44,45] Furthermore, this interplay of frailty and PerCI will better prepare us in caring for such patients in the next frontier beyond COVID-19. Future studies should focus on investigating the impact of frailty on the long-term outcomes of patients who developed PerCI.

### Strengths and Limitations

This study spanned numerous ICUs that enrolled patients across Australia and New Zealand, encompassing over 80% of patients critically unwell with COVID-19 during the first two years of the pandemic. The relatively larger sample of high-quality data increased the precision of our estimates. Additionally, we adjusted for appropriate confounders.

We must acknowledge a few limitations. The retrospective design of this study meant that data collection was reliant on existing datasets and medical records. In addition, there is a possibility of data coding inaccuracy, and without site-based auditing of diagnostic codes, we cannot be certain about the degree of misclassification if any, and what its effects are on our findings. Furthermore, as a retrospective registry-based study, it is only possible to highlight associations and no causal inferences can be drawn. We did not have any information regarding the number of patients that were referred for and denied ICU admission. In this study, the CFS was adopted in the assessment of frailty in ICUs across ANZ. Despite being an attractive tool to distinguish the different grades of frailty, the reliability of a single assessment tool may be inadequate, especially when it comes to justifying the rationing of medical treatment. Patients with COVID-19 admitted to the ICUs with an alternate diagnosis could have been missed. Additionally, many of the patients analysed had been transferred from another ICU, meaning that for these patients the true length of their ICU stay could not be determined, as this was not defined in the data set. Finally, the Australian and New Zealand healthcare systems have been very fortunate with the magnitude of COVID-19 infections at no point exceeding resource constraints, therefore the results may not be generalizable in all healthcare systems.

## Conclusion

This multicentre retrospective observational study of COVID-19 patients admitted to ICU in Australia and New Zealand revealed that hospital mortality was higher in patients with PerCI. The presence of frailty independently predicted hospital mortality in ICU patients, but the impact of frailty was no different in those who developed PerCI when compared to those who did not. This relationship between frailty and PerCI will help better cater to patient-centred treatment for such patients in the next frontier beyond COVID-19.

## Data Availability

All data produced in the present study are available upon reasonable request to the authors

## Acknowledgement

The authors and the ANZICS CORE management committee would like to thank clinicians, data collectors and researchers at the following contributing sites: Albury Base Hospital, Alfred Hospital, Alice Springs Hospital, Angliss Hospital, Armadale Health Service, Ashford Community Hospital, Auckland City Hospital CV, Auckland City Hospital DCCM, Austin Hospital, Ballarat Health Services, Bankstown-Lidcombe Hospital, Bathurst Base Hospital, Bendigo Health Care Group, Blacktown Hospital, Box Hill Hospital, Buderim Private Hospital, Bunbury Regional Hospital, Bundaberg Base Hospital, Caboolture Hospital, Cabrini Hospital, Cairns Hospital, Calvary Hospital (Canberra), Calvary Mater Newcastle, Calvary Wakefield Hospital (Adelaide), Campbelltown Hospital, Canberra Hospital, Casey Hospital, Central Gippsland Health Service, Christchurch Hospital, Coffs Harbour Health Campus, Concord Hospital (Sydney), Dandenong Hospital, Dubbo Base Hospital, Dunedin Hospital, Epworth Hospital (Richmond), Fairfield Hospital, Fiona Stanley Hospital, Flinders Medical Centre, Flinders Private Hospital, Footscray Hospital, Frankston Hospital, Gold Coast Private Hospital, Gold Coast University Hospital, Gosford Hospital, Gosford Private Hospital, Goulburn Base Hospital, Goulburn Valley Health, Grafton Base Hospital, Greenslopes Private Hospital, Griffith Base Hospital, Hawkes Bay Hospital, Hervey Bay Hospital, Hollywood Private Hospital, Holmesglen Private Hospital, Holy Spirit Northside Hospital, Hornsby Ku-ring-gai Hospital, Hutt Hospital, Ipswich Hospital, John Fawkner Hospital, John Hunter Hospital, Joondalup Health Campus, Kareena Private Hospital, Knox Private Hospital, Latrobe Regional Hospital, Launceston General Hospital, Lingard Private Hospital, Lismore Base Hospital, Liverpool Hospital, Logan Hospital, Lyell McEwin Hospital, Mackay Base Hospital, Macquarie University Private Hospital, Maitland Hospital, Maitland Private Hospital, Manning Rural Referral Hospital, Maroondah Hospital, Mater Adults Hospital (Brisbane), Mater Health Services North Queensland, Mater Private Hospital (Brisbane), Mater Private Hospital (Sydney), Melbourne Private Hospital, Middlemore Hospital, Mildura Base Hospital, Monash Medical Centre (Clayton), Mount Hospital, Mount Isa Hospital, Nelson Hospital, Nepean Hospital, Noosa Hospital, North Shore Hospital, North Shore Private Hospital, North West Regional Hospital (Burnie), Northeast Health Wangaratta, Northern Beaches Hospital, Norwest Private Hospital, Orange Base Hospital, Peninsula Private Hospital, Pindara Private Hospital, Port Macquarie Base Hospital, Prince of Wales Hospital (Sydney), Princess Alexandra Hospital, Queen Elizabeth II Jubilee Hospital, Redcliffe Hospital, Robina Hospital, Rockhampton Hospital, Rockingham General Hospital, Rotorua Hospital, Royal Adelaide Hospital, Royal Brisbane and Women’s Hospital, Royal Darwin Hospital, Royal Melbourne Hospital, Royal North Shore Hospital, Royal Perth Hospital, Royal Prince Alfred Hospital, Ryde Hospital & Community Health Services, Shoalhaven Hospital, Sir Charles Gairdner Hospital, South West Healthcare (Warrnambool), St Andrew’s Hospital (Adelaide), St Andrew’s Hospital Toowoomba, St Andrew’s Private Hospital (Ipswich), St Andrew’s War Memorial Hospital, St George Hospital (Sydney), St George Private Hospital (Sydney), St John of God (Berwick), St John Of God Health Care (Subiaco), St John of God Hospital (Bendigo), St John Of God Hospital (Geelong), St John Of God Hospital (Murdoch), St John of God Midland Public & Private, St Vincent’s Hospital (Melbourne), St Vincent’s Hospital (Sydney), St Vincent’s Hospital (Toowoomba), St Vincent’s Private Hospital Fitzroy, Sunnybank Hospital, Sunshine Coast University Hospital, Sunshine Coast University Private Hospital, Sunshine Hospital, Sutherland Hospital & Community Health Services, Sydney Adventist Hospital, Tamworth Base Hospital, Tauranga Hospital, The Chris O’Brien Lifehouse, The Northern Hospital, The Prince Charles Hospital, The Queen Elizabeth (Adelaide), The Townsville Hospital, The Valley Private Hospital, The Wesley Hospital, Timaru Hospital, Tweed Heads District Hospital, University Hospital Geelong, Wagga Wagga Base Hospital & District Health, Waikato Hospital, Warringal Private Hospital, Wellington Hospital, Werribee Mercy Hospital, Western District Health Service (Hamilton), Western Hospital (SA), Westmead Hospital, Whangarei Area Hospital, Northland Health Ltd, Wimmera Health Care Group (Horsham), Wollongong Hospital, and Wyong Hospital.

## Declarations

### Ethics approval and consent to participate

- All experimental protocols were approved by The Alfred Hospital Ethics Committee (Project No: 176/21) approved this study with a waiver of informed consent.

- ANZICS Centre for Outcome and Resource Evaluation Management Committee granted access to the ANZICS-APD in accordance with standing protocols.

- All methods were carried out in accordance with the relevant guidelines and regulations of the Declaration of Helsinki.

### Consent for publication

- Not applicable

### Availability of data and materials

- The datasets generated and/or analysed during the current study are not publicly available as these are from the ANZICS APD registry, but are available from the corresponding author upon reasonable request.

## Author contributions

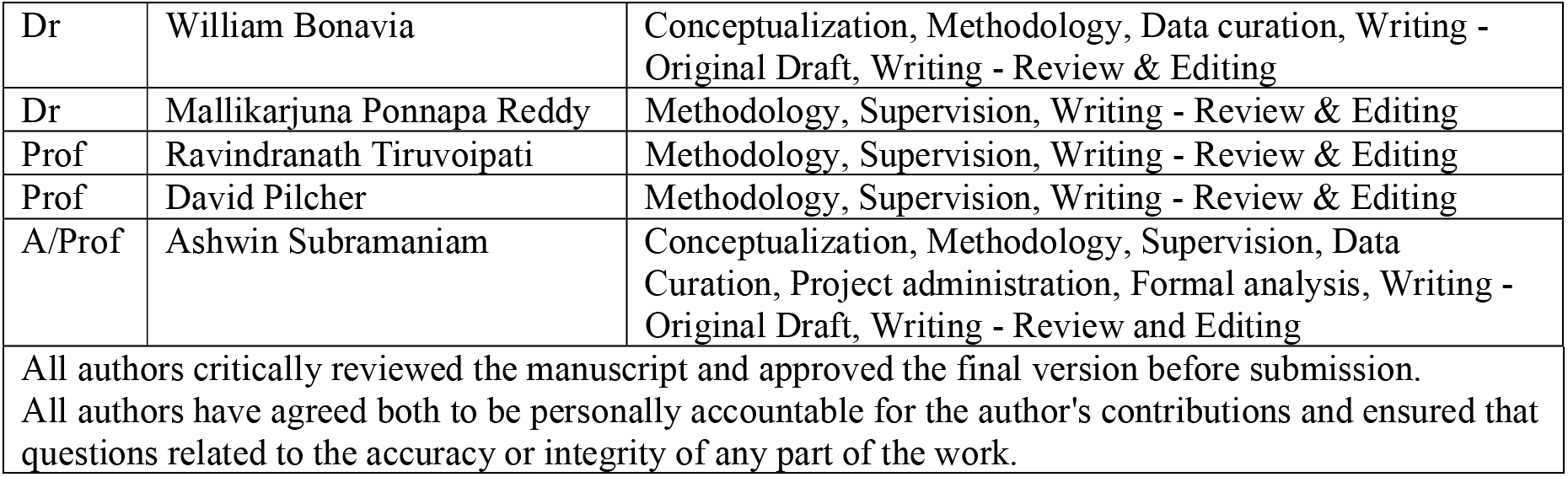

## Table and Figure Legends

**Supplementary Table 1:** Diagnostic codes and subcodes for patients included in the study between January 2020 and December 2021.

**Supplementary Table 2:** Missing data comparison for patients with and without the CFS scores.

**Supplementary Table 3a:** Baseline characteristics of patients with and without Persistent critical illness (PerCI), for clinical frailty scale (CFS) categories CFS-1-2 and CFS-3-4.

**Supplementary Table 3b:** Baseline characteristics of patients with and without Persistent critical illness (PerCI), for clinical frailty scale (CFS) categories CFS-5-6 and CFS-7-8.

**Supplementary Table 4:** Adjusted odds of hospital mortality in patients with and without persistent critical illness.

**Supplementary Figure 1:** Flow diagram demonstrating patient inclusion.

**Supplementary Figure 2:** Persistent Critical Illness (PerCI) predicting hospital mortality using unadjusted and adjusted (CFS adjusted for ANZROD and male sex) using AUROC (95%CI). Clinical Frailty Scale (CFS) is treated as a continuous variable.

**Supplementary Figure 3:** Proportion of patients with ICU length of stays and those who died in hospital, based on CFS categories.

## List of Abbreviations

ANZ: Australia and New Zealand
ANZICS: Australia and New Zealand Intensive Care Society
ANZROD: Australia and New Zealand risk of death
APACHE: Acute Physiology and Chronic Health Evaluation
APD: Adult Patient Database
AUROC: area under the receiver operating characteristic
COPD: chronic obstructive pulmonary disease
COVID-19: coronavirus disease 2019
CFS: Clinical Frailty Scale
HR: hazard ratio
ICU: intensive care unit
IQR: interquartile range
LOS: length of stay
MI: myocardial infarction
n: number
OR: odds ratio
PerCI: persistent critical illness
RRT: renal replacement therapy
SD: standard deviation

