## Supplemental Tables and Figures for "The impact of frailty on the outcomes of COVID-19 patients with persistent critical illness: A population-based cohort study"

**Supplementary Table 1:** Diagnostic codes and subcodes for patients included in the study between January 2020 and December 2021.

| | Diagnostic code in ANZICS-APD | Diagnostic subcode for patients without COVID-19 | Diagnostic subcode for patients with COVID-19* |
|---|---|---|---|
| Viral pneumonia | 213 | 213.01 | 213.02 |
| ARDS | 204 | 204.01 | 204.22 |

ANZICS – Australia New Zealand Intensive Care Society; APD – adult patient database; ARDS – acute respiratory distress syndrome

* The patients were considered highly likely to be positive for COVID-19 ("Suspected or confirmed pandemic infection"), as ANZICS does not have this information but infers it.

**Supplementary Table 2:** Missing data comparison for patients with and without available CFS scores.

| Variable | Frailty data present (n=3064) | Missing frailty data (n=645) | p-value |
|---|---|---|---|
| PerCI, n (%) | 847 (27.6%) | 134 (20.9%) | **<0.001** |
| Indigenous, n (%) | 78 (2.5%) | 10 (1.6%) | 0.32 |
| Male sex, n (%) | 1879 (56.2%) | 382 (59.6%) | 0.41 |
| Age (years) (median [IQR]) | 56.9 [44.7, 68.2] | 59.0 [48.3, 69.9] | **0.007** |
| **Hospital admission source, n (%)** | | | |
| - Home | 2473 (80.7%) | 539 (84.1%) | **0.012** |
| - Other acute hospital (not ICU/ED) | 129 (4.2%) | 16 (2.5%) | |
| - Nursing home or chronic care | 15 (0.5%) | 8 (1.2%) | |
| - Other hospital ICU | 257 (8.4%) | 38 (5.9%) | |
| - Other hospital Emergency department | 151 (4.9%) | 28 (4.4%) | |
| - Rehabilitation | 3 (0.1%) | 2 (0.3%) | |
| **ICU admission source, n (%)** | | | |
| - Emergency department | 1191 (38.9%) | 280 (43.7%) | **<0.001** |
| - Ward | 1483 (48.4%) | 278 (43.4%) | |
| - Other hospital | 103 (3.4%) | 33 (5.1%) | |
| - Other hospital ICU | 272 (8.9%) | 36 (5.6%) | |
| - Operating theatre / Recovery | 1 (0.0%) | 0 (0) | |
| - Direct admit | 14 (0.5%) | 14 (2.2%) | |
| **Documented co-morbidities, n (%)** | | | |
| - Chronic respiratory condition | 200 (6.5%) | 15 (2.3%) | **<0.001** |
| - Chronic cardiovascular condition | 179 (5.8%) | 19 (3.0%) | **0.003** |
| - Chronic renal failure | 74 (2.4%) | 6 (0.9%) | **0.019** |
| - Chronic liver disease | 21 (0.7%) | 6 (0.9%) | 0.50 |
| - Diabetes mellitus | 911 (29.7%) | 89 (13.9%) | **<0.001** |
| - Immune suppressive therapy | 142 (4.6%) | 27 (4.2%) | 0.64 |
| - Lymphoma | 13 (0.4%) | 1 (0.2%) | 0.31 |
| - Leukaemia | 26 (0.8%) | 6 (0.9%) | 0.83 |
| - Metastatic cancer | 24 (0.8%) | 4 (0.6%) | 0.67 |
| - Delirium, n (%) | 259 (8.5%) | 8 (1.2%) | **<0.001** |
| **Organ failure score** | | | |
| - APACHE III score (mean [SD]) | 50.0 [20.0] | 50.4 [21.7] | 0.47 |
| - ANZROD (%) (mean [SD]) | 9.5 [12.2] | 10.1 [12.1] | 0.61 |
| ICU admission post MET call, n (%) | 1105 (36.3%) | 182 (28.4%) | **<0.001** |
| Cardiac arrest, n (%) | 6 (0.2%) | 0 (0) | **0.004** |
| Treatment limitation, n (%) | 255 (8.4%) | 55 (8.6%) | 0.40 |
| Pre-ICU (days) (median [IQR]) | 12.4 [4.0, 56.8] | 9.5 [3.7, 63.6] | 0.31 |
| **ICU supports** | | | |
| Mechanical ventilation (MV), n (%) | 1306 (43.1%) | 195 (32.0%) | **<0.001** |
| Non-invasive ventilation (NIV), n (%) | 1260 (41.8%) | 203 (33.4%) | **<0.001** |
| Vasopressor and inotropes, n (%) | 1188 (39.2%) | 200 (32.8%) | **0.003** |
| Renal replacement therapy, n (%) | 180 (6.0%) | 33 (5.5%) | 0.63 |
| ECMO, n (%) | 104 (3.5%) | 9 (1.5%) | **0.011** |
| Tracheostomy, n (%) | 186 (6.2%) | 13 (2.2%) | **<0.001** |

CFS – clinical frailty scale, SD – standard deviation, IQR. – interquartile range, MET – medical emergency team, APACHE - Acute Physiology and Chronic Health Evaluation, ED – emergency department, ICU – intensive care unit, ROD – risk of death, ANZROD – Australia New Zealand risk of death, PerCI - Persistent critical illness

**Supplementary Table 3a:** Baseline characteristics of patients with and without Persistent critical illness (PerCI), for clinical frailty scale (CFS) categories CFS-1-2 and CFS-3-4.

| Variable | CFS-1-2 (n=1123) | | | CFS-3-4 (n=1575) | | |
|---|---|---|---|---|---|---|
| | Patients without PerCI (n=835) | Patients with PerCI (n=288) | p-value | Patients without PerCI (n=1109) | Patients with PerCI (n=466) | p-value |
| Male sex, n (%) | 523 (62.6%) | 203 (70.5%) | **0.012** | 656 (59.2%) | 295 (63.3%) | 0.12 |
| Indigenous status, n (%) | 24 (2.9%) | 8 (2.8%) | 0.92 | 32 (2.9%) | 9 (1.9%) | 0.46 |
| Age, (years), median (IQR) | 46.5 (34.1, 56.6) | 54.0 (43.5, 63.9) | **<0.001** | 59.1 (47.5, 69.3) | 61.7 (51.7, 69.8) | **0.010** |
| **Hospital admission source, n (%)** | | | | | | |
| - Home | 715 (85.6%) | 191 (66.3%) | **<0.001** | 950 (85.7%) | 337 (72.3%) | **<0.001** |
| - Other acute hospital | 103 (12.4%) | 90 (31.3%) | | 148 (13.3%) | 126 (26.9%) | |
| - Nursing home or chronic care | 0 (0) | 1 (0.3%) | | 1 (0.1%) | 0 (0) | |
| - Rehabilitation | 0 (0) | 0 (0) | | 2 (0.2%) | 0 (0) | |
| - Missing | 17 (2.0%) | 6 (2.1%) | | 8 (0.7%) | 4 (0.9%) | |
| **ICU admission source, n (%)** | | | | | | |
| - Emergency department (ED) | 355 (42.5%) | 93 (32.3%) | **<0.001** | 459 (41.4%) | 138 (29.6%) | **<0.001** |
| - Ward | 411 (49.2%) | 116 (40.3%) | | 340 (45.1%) | 40 (43.0%) | |
| - Other hospital (ED and ICU) | 62 (7.4%) | 77 (26.7%) | | 179 (23.7%) | 30 (32.3%) | |
| - Operating theatre / Recovery | 1 (0.1%) | 0 (0) | | 0 (0) | 0 (0) | |
| - Direct admit | 6 (0.7%) | 2 (0.7%) | | 2 (0.2%) | 2 (0.4%) | |
| **Documented co-morbidities, n (%)** | | | | | | |
| - Chronic respiratory condition | 25 (3.0%) | 7 (2.4%) | 0.62 | 72 (6.5%) | 17 (3.6%) | **<0.001** |
| - Chronic CVS condition | 18 (2.2%) | 6 (2.1%) | 0.94 | 71 (6.4%) | 16 (3.4%) | **0.019** |
| - Chronic renal failure | 3 (0.4%) | 1 (0.3%) | 0.98 | 30 (2.7%) | 10 (2.1%) | 0.52 |
| - Chronic liver disease | 0 (0) | 1 (0.3%) | 0.09 | 7 (0.6%) | 4 (0.9%) | 0.62 |
| - Diabetes mellitus | 138 (16.9%) | 63 (22.1%) | 0.05 | 375 (35.6%) | 181 (41.5%) | **0.031** |
| - Immune suppressive therapy | 12 (1.4%) | 30 (11.0%) | 0.45 | 58 (5.2%) | 26 (5.6%) | 0.78 |
| - Lymphoma | 0 (0) | 0 (0) | - | 5 (0.5%) | 3 (0.6%) | 0.62 |
| - Leukaemia | 1 (0.1%) | 1 (0.3%) | 0.43 | 8 (0.7%) | 5 (1.1%) | 0.48 |
| - Metastatic cancer | 2 (0.2%) | 0 (0) | 0.41 | 9 (0.8%) | 2 (0.4%) | 0.41 |
| - Obese (BMI ≥ 30 $kg.m^{-2}$) | 410 (49.1%) | 100 (34.7%) | **<0.001** | 440 (39.7%) | 161 (34.5%) | 0.11 |
| - Delirium | 16 (1.9%) | 44 (15.3%) | **<0.001** | 279 (25.2%) | 124 (26.6%) | **<0.001** |
| - Pregnancy status | 31 (3.7%) | 6 (2.1%) | **<0.001** | 31 (2.8%) | 4 (0.9%) | **0.004** |
| Pre-ICU (days) (median [IQR]) | 13.6 (4.6, 54.4) | 6.7 (0.9, 36.8) | **<0.001** | 13.5 (4.6, 59.3) | 9.9 (1.6, 57.9) | **0.004** |
| ICU discharge delay, (median [IQR]) | 4.0 (2.2, 7.2) | 5.0 (2.9, 11.4) | **<0.001** | 4.0 (1.9, 7.1) | 4.5 (1.5, 8.2) | 0.48 |
| **Organ failure scores** | | | | | | |
| - APACHE III (mean [SD]) | 39.6 (16.6) | 53.2 (18.4) | **<0.001** | 49.9 (11.9) | 57.3 (18.1) | **<0.001** |
| - ANZROD (%) (mean [SD]) | 4.6 (6.0) | 9.1 (10.6) | **<0.001** | 9.3 (11.9) | 11.1 (10.7) | **<0.001** |
| ICU admission post MET call, n (%) | 282 (34.0%) | 88 (30.9%) | 0.34 | 432 (39.3%) | 164 (35.5%) | 0.16 |
| Treatment limitations | 7 (0.8%) | 6 (2.1%) | 0.13 | 86 (7.8%) | 19 (4.1%) | **0.026** |
| Cardiac arrest, n (%) | 0 (0) | 0 (0) | - | 4 (0.4%) | 0 (0) | 0.39 |
| **ICU Supports**, n (%) | | | | | | |
| Mechanical ventilation | 168/823 (20.4%) | 247/286 (86.4%) | **<0.001** | 365/1097 (33.3%) | 402/465 (86.5%) | **<0.001** |
| Non-invasive ventilation | 270/823 (32.8%) | 111/282 (39.4%) | **0.046** | 476/1093 (43.5%) | 234/461 (50.8%) | **0.009** |
| Vasopressor and inotropes | 130/824 (15.8%) | 216/285 (75.8%) | **<0.001** | 339/1097 (30.9%) | 369/464 (79.5%) | **<0.001** |
| Renal replacement therapy | 6/822 (0.7%) | 39/282 (13.8%) | **<0.001** | 29/1091 (2.7%) | 85/461 (18.4%) | **<0.001** |
| ECMO | 6/822 (0.7%) | 45/282 (16.0%) | **<0.001** | 6/1092 (0.5%) | 45/461 (9.8%) | **<0.001** |
| Tracheostomy | 3/822 (0.4%) | 55/281 (19.6%) | **<0.001** | 3/1090 (0.3%) | 111/458 (24.2%) | **<0.001** |

CFS – clinical frailty scale, SD – standard deviation, IQR. – interquartile range, BMI – body mass index, MET – medical emergency team, APACHE - Acute Physiology and Chronic Health Evaluation, ED – emergency department, ICU – intensive care unit, ANZROD – Australia New Zealand risk of death. ECMO – extracorporeal membrane oxygenation

**Supplementary Table 3b:** Baseline characteristics of patients with and without Persistent critical illness (PerCI), for clinical frailty scale (CFS) categories CFS-5-6 and CFS-7-8.

| **Variable** | **CFS-5-6 (n=299)** | | | **CFS-7-8 (n=67)** | | |
|---|---|---|---|---|---|---|
| | **Patients without PerCI (n=219)** | **Patients with PerCI (n=80)** | **p-value** | **Patients without PerCI (n=54)** | **Patients with PerCI (n=13)** | **p-value** |
| Male sex, n (%) | 105 (47.9%) | 50 (62.5%) | **0.026** | 36 (66.7%) | 11 (84.6%) | 0.20 |
| Indigenous status, n (%) | 1 (0.5%) | 0 (0) | 0.75 | 4 (7.4%) | 0 (0) | 0.31 |
| Age, (years), median (IQR) | 72.8 (64.5, 81.6) | 68.3 (58.0, 75.4) | **<0.001** | 68.7 (55.8, 78.5) | 62.1 (52.1, 70.3) | 0.20 |
| **Hospital admission source, n (%)** | | | | | | |
| - Home | 185 (84.5%) | 48 (60.0%) | **<0.001** | 41 (75.9%) | 6 (46.2%) | **0.003** |
| - Other acute hospital | 27 (12.3%) | 29 (36.4%) | | 8 (14.9%) | 7 (53.9%) | |
| - Nursing home or chronic care | 7 (3.2%) | 2 (2.5%) | | 4 (7.4%) | 0 (0) | |
| - Rehabilitation | 0 (0) | 0 (0) | | 1 (1.9%) | 0 (0) | |
| - Missing | 0 (0) | 1 (1.3%) | | 0 (0) | 0 (0) | |
| **ICU admission source, n (%)** | | | | | | |
| - Emergency department (ED) | 99 (45.2%) | 21 (26.3%) | **<0.001** | 24 (44.4%) | 2 (15.4%) | **0.018** |
| - Ward | 111 (50.7%) | 35 (43.8%) | | 23 (42.6%) | 5 (38.5%) | |
| - Other hospital (ED and ICU) | 8 (3.6%) | 24 (30.1%) | | 6 (11.1%) | 6 (46.2%) | |
| - Operating theatre / Recovery | 0 (0) | 0 (0) | | 0 (0) | 0 (0) | |
| - Direct admit | 1 (0.5%) | 0 (0) | | 1 (1.9%) | 0 (0) | |
| **Documented co-morbidities, n (%)** | | | | | | |
| - Chronic respiratory condition | 48 (21.9%) | 9 (11.3%) | **0.038** | 18 (33.3%) | 4 (30.8%) | 0.86 |
| - Chronic CVS condition | 45 (20.5%) | 7 (8.8%) | **0.017** | 15 (27.8%) | 1 (7.7%) | 0.13 |
| - Chronic renal failure | 24 (11.0%) | 2 (2.5%) | **0.022** | 4 (7.4%) | 0 (0) | 0.31 |
| - Chronic liver disease | 5 (2.3%) | 2 (2.5%) | 0.91 | 2 (3.7%) | 0 (0) | 0.48 |
| - Diabetes mellitus | 97 (45.8%) | 34 (43.6%) | 0.74 | 22 (41.5%) | 1 (7.7%) | **0.022** |
| - Immune suppressive therapy | 25 (11.4%) | 9 (11.3%) | 0.97 | 5 (9.3%) | 1 (7.7%) | **0.040** |
| - Lymphoma | 3 (1.4%) | 1 (1.3%) | 0.94 | 0 (0) | 1 (7.7%) | 0.62 |
| - Leukaemia | 6 (2.7%) | 2 (2.5%) | 0.91 | 3 (5.6%) | 0 (0) | 0.39 |
| - Metastatic cancer | 7 (3.2%) | 1 (1.3%) | 0.36 | 2 (3.7%) | 1 (7.7%) | 0.53 |
| - Obese (BMI ≥ 30 $kg.m^{-2}$) | 55 (25.1%) | 32 (40.0%) | **0.028** | 19 (35.2%) | 7 (53.8%) | 0.41 |
| - Delirium | 23 (10.5%) | 18 (22.5%) | **0.006** | 7 (13.0%) | 3 (23.1%) | 0.50 |
| - Pregnancy status | 0 (0) | 0 (0) | **-** | 0 (0) | 0 (0) | **-** |
| Pre-ICU (days) (median [IQR]) | 17.9 (6.4, 75.9) | 5.5 (0.1, 69.4) | **<0.001** | 15.6 (5.5, 88.9) | 13.6 (0.1, 61.6) | 0.20 |
| ICU Discharge delay | 4.2 (1.3, 8.8) | 4.9 (1.9, 9.9) | 0.57 | 4.8 (2.0, 11.5) | 0 (0, 2.9) | **0.006** |
| **Organ failure scores** | | | | | | |
| - APACHE III (mean [SD]) | 62.1 (20.6) | 62.8 (20.3) | 0.61 | 62.5 (24.0) | 66.5 (26.8) | 0.80 |
| - ANZROD (%) (mean [SD]) | 20.1 (18.4) | 16.4 (14.6) | 0.10 | 23.8 (24.0) | 19.6 (17.3) | 0.86 |
| ICU admission post MET call, n (%) | 91 (41.6%) | 25 (32.1%) | 0.14 | 18 (33.3%) | 5 (38.5%) | 0.73 |
| Treatment limitations | 94 (43.0%) | 11 (13.8%) | **<0.001** | 30 (55.6%) | 2 (14.4%) | **0.016** |
| Cardiac arrest, n (%) | 0 (0) | 0 (0) | - | 1 (1.9%) | 1 (7.7%) | 0.54 |
| **ICU Supports**, n (%) | | | | | | |
| Mechanical ventilation | 37/213 (17.4%) | 61/80 (76.3%) | **<0.001** | 14/53 (26.4%) | 12/13 (92.3%) | **<0.001** |
| Non-invasive ventilation | 102/213 (47.9%) | 38/77 (49.4%) | 0.83 | 27/54 (50.0%) | 2/13 (15.4%) | **0.024** |
| Vasopressor and inotropes | 48/214 (22.4%) | 58/80 (72.5%) | **<0.001** | 17/53 (32.1%) | 11/13 (84.6%) | **<0.001** |
| Renal replacement therapy | 9/213 (4.2%) | 8/77 (10.4%) | **0.048** | 1/53 (1.9%) | 3/413 (23.1%) | **0.004** |
| ECMO | 0/213 (0) | 2/77 (2.6%) | **0.018** | 0 (0) | 0 (0) | **-** |
| Tracheostomy | 2/213 (0.9%) | 11/77 (14.3%) | **<0.001** | 0 (0) | 1/13 (7.7%) | **0.042** |

CFS – clinical frailty scale, SD – standard deviation, IQR. – interquartile range, BMI – body mass index, MET – medical emergency team, APACHE - Acute Physiology and Chronic Health Evaluation, ED – emergency department, ICU – intensive care unit, ANZROD – Australia New Zealand risk of death. ECMO – extracorporeal membrane oxygenation

**Supplementary Table 4:** Adjusted odds of hospital mortality in patients with and without persistent critical illness. The graphical version is presented in Figure 1.

| **Predictor** | **Patients without Persistent Critical illness** | | **Patients with Persistent Critical illness** | |
|---|---|---|---|---|
| | **OR (95% CI)** | **p-value** | **OR (95% CI)** | **p-value** |
| **Logistic Regression Model 1 – Frailty assessed using CFS categories** | | | | |
| CFS 1-2 | Reference | | Reference | |
| CFS 3-4 | 3.25 (1.86-5.66) | **<0.001** | 2.09 (1.44-3.04) | **<0.001** |
| CFS 5-6 | 5.54 (2.88-10.66) | **<0.001** | 1.87 (1.03-3.40) | **0.041** |
| CFS 7-8 | 11.02 (4.52-26.85) | **<0.001** | 7.01 (1.94-25.26) | **0.003** |
| Male sex | 1.77 (1.20-2.59) | **<0.001** | 1.31 (0.93-1.85) | 0.12 |
| ANZROD | 1.08 (1.07-1.10) | **<0.001** | 1.05 (1.04-1.07) | **<0.001** |
| **Logistic Regression Model 2 – Frailty assessed using CFS as a continuous variable** | | | | |
| CFS | 1.46 (1.29-1.64) | **<0.001** | 1.30 (1.14-1.49) | **<0.001** |
| ANZROD | 1.08 (1.07-1.10) | **<0.001** | 1.05 (1.04-1.07) | **<0.001** |
| Male sex | 1.82 (1.24-2.67) | **0.002** | 1.32 (0.94-1.86) | 0.11 |
| CFS = clinical frailty scale, ANZROD = Australia and New Zealand risk of death | | | | |

**Supplementary Figure 1:** Flow diagram demonstrating patient inclusion.

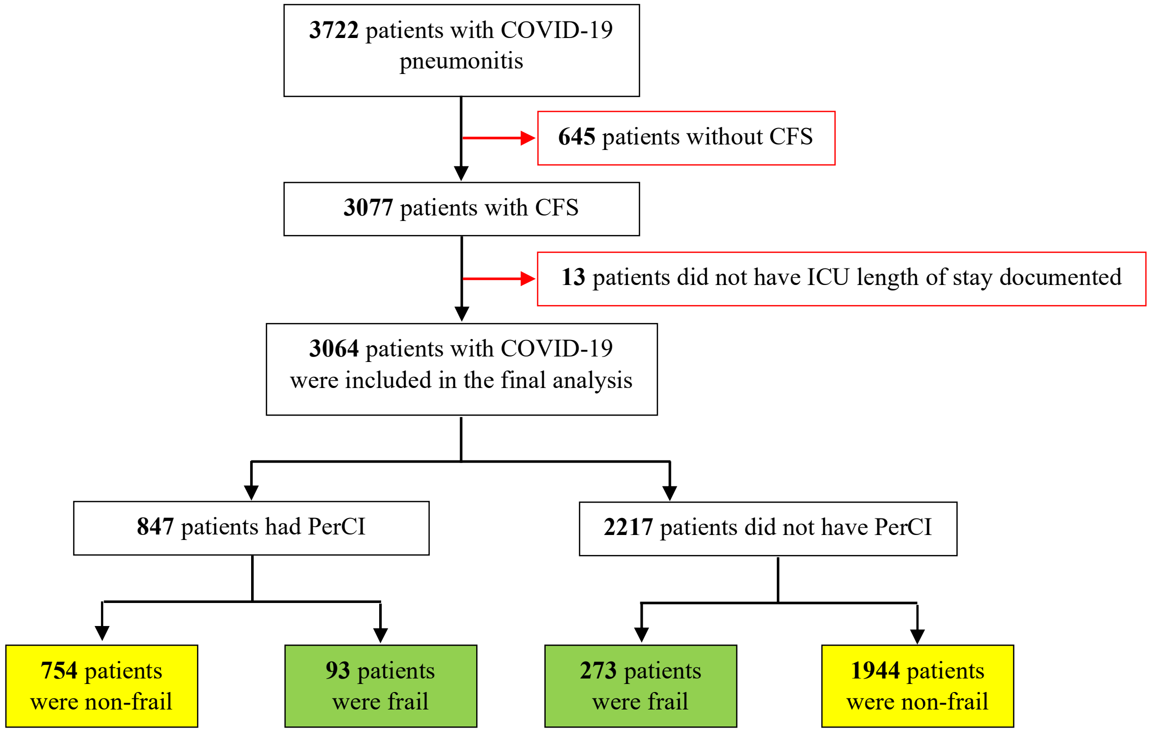


CFS – clinical frailty scale, PerCI - Persistent critical illness
Non-frail (CFS 1-4), Frail (CFS 5-8)

**Supplementary Figure 2:** The relationship between frailty and mortality in those with and without PerCI, before and after adjusting for ANZROD and male sex, using AUROC (95%CI). The Clinical Frailty Scale (CFS) was treated as a continuous variable.

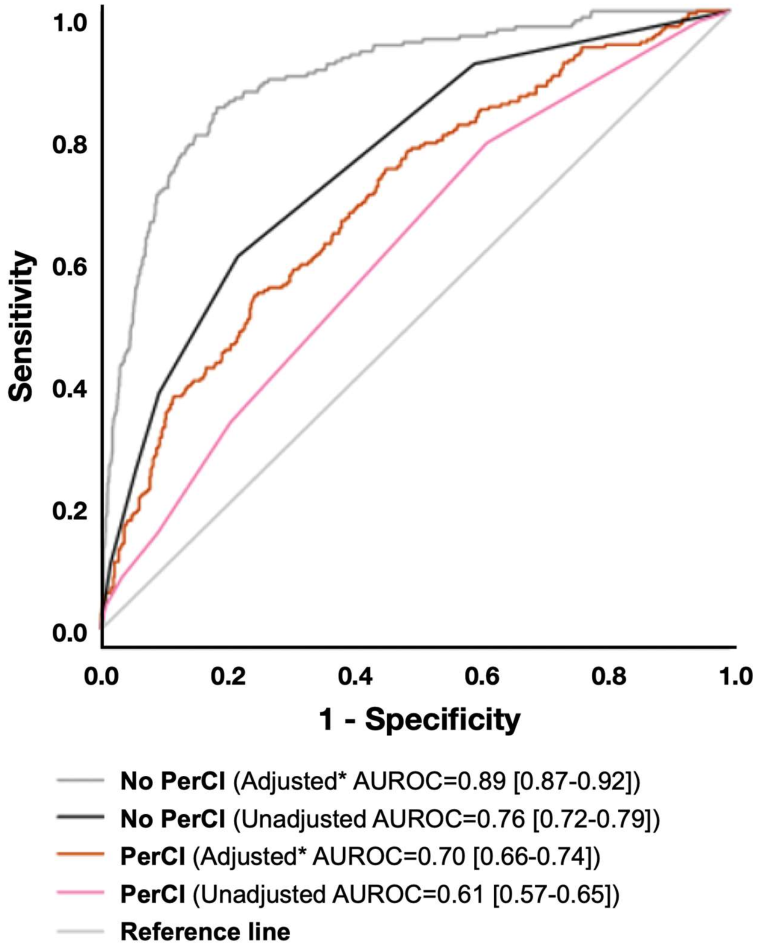


Comparison between unadjusted and adjusted PerCI vs. no PerCI are both <0.001
* Adjusted CFS for ANZROD and male sex

**Supplementary Figure 3:** Proportion of patients with ICU length of stays and those who died in hospital, based on CFS categories. *Error bars are standard errors of the mean.

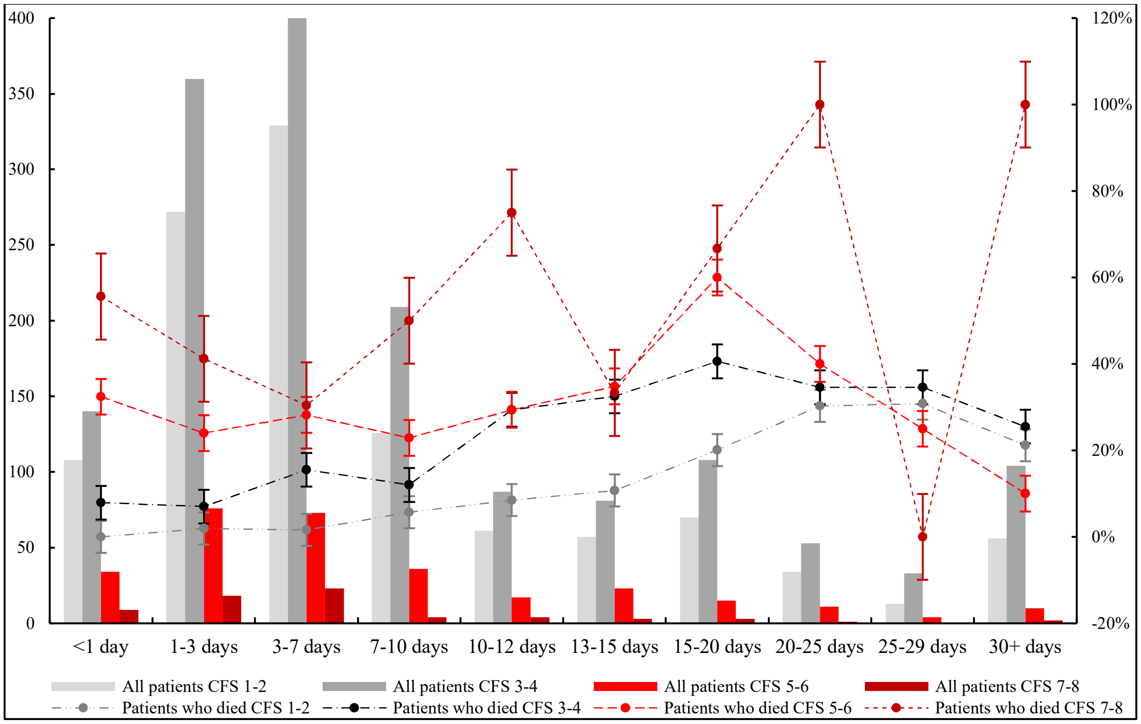
